# Polygenic risk score analysis reveals shared genetic burden between epilepsy and psychiatric comorbidities

**DOI:** 10.1101/2023.07.04.23292071

**Authors:** C. Campbell, D. Lewis-Smith, C. Leu, H. Martins, S. Wolking, R. Krause, T. O’Brien, G. Sill, F. Zara, B. Koeleman, C. Depondt, A. Marson, H. Stefánnson, K. Stefánnson, J. Craig, MR. Johnson, P. Striano, A. Jorgensen, H. Lerche, N. Delanty, The EpiPGX Consortium, S.M. Sisodiya, R. H. Thomas, G. L. Cavalleri

## Abstract

**Background:** One in three people with epilepsy experiences psychiatric comorbidity, with higher rates in people with drug-resistant epilepsy. Despite their high heritabilities, finding genetic links between epilepsy and psychiatric disorders has proven difficult. We used polygenic risk scoring (PRS) to test whether people with epilepsy have an increased polygenic burden of common genetic variants for depression, anxiety, psychosis, and attention deficit/hyperactivity disorder (ADHD), and examined whether such polygenic burden influences the response to pharmacological treatment of epilepsy.

**Methods:** Phenotype data in the UK Biobank were assessed to identify people with 1) epilepsy (n=8 488), 2) depression (n=143 440), 3) psychosis (n=2 357), 4) ADHD (n=89), and 5) anxiety (n=18 222. Using genotype data and restricting to Caucasian-ancestry samples (n=409 634), PRS for each psychiatric trait were calculated and multinomial regression was used to compare 1) population controls, 2) people with epilepsy and no psychiatric illness, 3) people with epilepsy and the psychiatric trait of interest, and 4) people with the psychiatric trait of interest and no epilepsy. Fixed-effect meta-analysis was used to compare psychiatric PRS in drug-resistant and drug-responsive epilepsy samples from the UK Biobank (n=1 640) and the EpiPGX consortium (n=3 449).

**Results:** After correction for multiple testing, people with epilepsy showed elevated PRS for depression (p<2 x10^−16^), psychosis (p=0.04) and ADHD (p<0.001). Patients with drug-resistant epilepsy had an increased PRS for psychosis (p=0.002) and depression (p=0.0004) relative to responsive cases.

**Conclusions:** We present evidence that the common genetic basis of epilepsy overlaps with that of psychiatric conditions which are frequently comorbid in people with epilepsy. Common genetic variants that drive psychiatric illness are enriched in people with drug-resistant epilepsy. These results further our understanding of the genetic architecture of epilepsy and suggest a potential future role for polygenic interpretation of common variants in prognostic stratification, both for seizure-treatment outcomes and non-seizure comorbidities.

## Introduction

Epilepsy is a common neurological disorder affecting nearly 70 million people worldwide^1, 2^. The aetiology of epilepsy is complex, with both common and rare genetic variants playing a role in disease susceptibility, as well as environmental factors^3, 4^. Differences in the underlying aetiology of epilepsy can affect disease presentation, such as seizure localisation, age of seizure onset, or developmental impairment^4^. Anti-seizure medicines (ASM), either alone or in combination, are effective in controlling seizures in 64–70% of people with epilepsy^5, 6^. Among the many adverse consequences of drug-resistant epilepsy is a higher rate of psychiatric comorbidities.

Psychiatric comorbidities are commonly seen in people with epilepsy, with a lifetime prevalence of 35%^7, 8^. Affective disorders are the most common psychiatric comorbidity in adults, and ADHD in children^9^. People with epilepsy, as well as their immediate family members, are at an increased risk of a variety of psychiatric disorders such as ADHD, anxiety, depression and schizophrenia relative to the general population^7, 9, 10^. Psychiatric comorbidities are particularly enriched among people with drug-resistant seizures^11^. A small number of rare genetic conditions such as 22q11 microdeletion syndrome^12^ and *PCDH19*-associated epilepsy^13^ demonstrate propensity to both seizures and psychopathology, however recent studies did not provide evidence for a genetic link between common epilepsies and psychiatric disorders^14, 15^.

Linkage-disequilibrium score regression (LDSC) is a method which compares test statistics (beta coefficients or odds ratios) from genome-wide association studies (GWAS) to patterns of linkage disequilibrium to quantify signal related to polygenicity, distinct from confounding factors (e.g. genetic ancestry)^16^. LDSC can be used to compare the polygenic basis of complex conditions. This approach has successfully correlated common genetic risk between many psychiatric disorders which are known to co-occur, such as ADHD, bipolar disorder, major depressive disorder and schizophrenia^15^. However, thus far, LDSC analysis has been unable to find genetic correlations between epilepsy, or any epilepsy subtypes, and any psychiatric disorder^15, 17^. This could be due to lack of study power, and/or that LDSC is less suited to the genetic architecture of epilepsy than it is to that of other traits^18^.

Polygenic risk scoring (PRS) is a method which quantifies a person’s common genetic burden for a trait or condition, derived from GWAS results^19^. Epilepsy PRS has been demonstrated to be capable of distinguishing people with epilepsy from controls, however this observation was stronger in datasets developed from patients attending tertiary epilepsy referral centres than in less selective population-wide datasets, such as the UK Biobank (UKB)^20^. PRS analyses can also be used to inspect genetic correlations between datasets and are more sensitive than alternative methods (e.g., LDSC) when inspecting genetic correlation between traits or cohorts where the proportion of heritability conferred by the top-associated loci is unknown^21, 22^. For example, PRS derived from a schizophrenia GWAS have been shown to predict bipolar disorder in case:control analyses^23^. Epilepsy PRS have been found be pleiotropic across several brain-related phenotypes, including neuroticism, educational attainment and three broad self-reported statements indicating symptoms of anxiety or depression^24^.

Here we aim to leverage the cohort size and diverse phenotypic data of the UKB to test the hypothesis that PRS for a variety of psychiatric disorders are elevated in people with epilepsy. The traits we examined were ADHD, anxiety, depression, and schizophrenia. These traits were chosen as they are frequently comorbid in people with epilepsy and had available GWAS summary statistics (for PRS calculation) with no sample overlap with the UKB. We compared the PRS for the above psychiatric traits in people with epilepsy, both with and without the trait of interest, as well as to that of the general population, and cases of psychiatric illness without a diagnosis of epilepsy. We then aimed to compare the polygenic burden for these same psychiatric illnesses in cases of drug-resistant and drug-responsive epilepsy, using data from the UKB and the EpiPGX consortium.

## Materials and methods

### Dataset

All genotype and phenotype data were from the UKB^25^. Data access was approved under UKB project proposal 35124. After phenotyping, all samples were screened for Caucasian ancestry using UKB field 220066 (genetic ethnic grouping). For EpiPGX samples, ethical approval was obtained from the ethics committee of each referral centre. Informed consent was obtained from all patients or where applicable, from their legal guardians, during routine clinic attendance.

### UKB phenotyping & case-control assignment

We used the phenotypic information provided by UKB to classify samples non-exclusively into those from individuals with the following conditions: epilepsy, psychosis, depression, anxiety, and ADHD. For each of the traits studied, samples were divided into the following groups: 1) epilepsy only, 2) epilepsy AND psychiatric phenotype, 3) psychiatric phenotype only, and 4) controls.

#### Identification of people with epilepsy in the UKB

We identified people predicted to have active or previous epilepsy from the UKB cohort recruited at ages 40–69 years, between 2007 and 2010, using data released between 2^nd^ July and 25^th^ September 2020 (502 493 individuals). We searched self-reported data, inpatient hospital episode statistics (HES), death certificates diagnostic data for “epilepsy” or “status epilepticus”. We also searched the primary care diagnostic codes of the 230 090 individuals for whom these records were available. We searched these data sources independently to allow appraisal of the evidence of affectation rather than relying on the provided UKB epilepsy diagnosis (data fields 131048 and 13049, which pool diagnoses mapped to ICD-10 code G40 (epilepsy) from these sources). This allowed us to exclude individuals without sufficient evidence of epilepsy, such as those with seizures due solely to an unspecified disorder or condition that is not considered epilepsy, and to identify further cases according to our criteria that may have been overlooked in the UKB provided cohort (Table 1). While the use of ASM data can increase the sensitivity of case identification from administrative records^26^, the positive predictive value of ASM for a diagnosis of epilepsy is poor if not combined with positive diagnostic data^27^. Consequently, we elected not to use the presence of self-declared or prescribed ASM to predict epilepsy affectation.

**Table 1:** Inclusion criteria for UK Biobank epilepsy cohort definition.

| **UK Biobank epilepsy cohort definition** |  |
| --- | --- |
| **Data** | **Criteria** |
| HES inpatient ICD-9 and ICD-10 codes | ICD-10: F803, G400-G419, P90X |
|  | ICD-9: 3450-3459, 7790 |
| Self-reported diagnosis | Epilepsy (value 1264) |
| Primary care Read 2 and Read 3 diagnostic codes | Codes indicating previous or current epilepsy, status epilepticus, epilepsy treatment, epilepsy monitoring, and epilepsy outcomes were considered sufficient.<br>(Codes used for indicating seizure types, alcohol- and other substance associated seizures, non-epileptic dissociative seizures, EEG findings, antiseizure medication, a family history or relationship with someone who has epilepsy, or a referral to an epilepsy clinic were considered insufficient) |
| Death certification | Epilepsy or status epilepticus listed as primary or contributing cause of death |
| **Drug resistant group definition** |  |
| HES inpatient diagnostic codes and dates | Primary cause of admission > 10 years after earliest diagnosis of epilepsy or status epilepticus * |
| HES inpatient diagnostic codes and dates | Primary cause of multiple admissions with the most recent > 5 years after the first and with a mean interval between admissions < 5 years |
| OPCS-3 and OPCS-4 procedural codes | Surgical treatment suggestive of drug-resistance identified by manual review after screening for single episodes coded with brain or vagal nerve surgery and involving EEG, electrocorticography or a neurostimulator |
| OPCS-3 and OPCS-4 procedural codes | $\geq 2$ EEGs performed > 5 years after earliest diagnosis * |
| Self-reported medication | $\geq 3$ ASM at time of enrolment |
| Primary care Read 2 and Read 3 prescription codes and dates | $\geq 3$ ASM prescribed within 3 months of most recent prescription |
| HES inpatient diagnostic codes and dates, and Primary care Read 2 and Read 3 prescription codes and dates | Primary cause of admission or commencement of a 4<sup>th</sup> ASM after being prescribed 3 different ASM $\geq 6$ months |
| Death certification | Primary cause of death is epilepsy, status epilepticus, or sudden unexpected death in epilepsy |
| **Responsive group definition** |  |
| Diagnosis of epilepsy before 2000/01/01, * and no EEG or admission with epilepsy or status epilepticus as a primary cause since 2005/01/01, with either: |  |
| A) No self-reported ASM at time of enrolment and since 2005/01/01 no primary care prescription for ASM |  |
| B) Driver at enrolment and complete follow-up data collection without evidence of commencing a new ASM since 2005/01/01 |  |

**Table legend:** \* Earliest date of diagnosis was identified from HES inpatient, primary care, and pooled UKB earliest diagnostic dates. HES = Hospital Episode Statistics, ICD = International Classification of Disease, OPCS = Office of Population Censuses and Surveys Classification of Interventions and Procedures, ASM = anti-seizure medication, Acetazolamide, gabapentin, and pregabalin are not included as ASM in this manuscript. Individuals meeting any criteria above were allocated to the respective cohort, except for those meeting inclusion criteria for both the Resistant and Responsive groups who were excluded from both strata.

The codes used for epilepsy filters can be found in the supplemental materials (Supplementary Tables S1-5).

#### Identification of people with ADHD, anxiety, depression or psychosis in the UKB

For anxiety^28^, depression^29^ and psychosis^30^ we used previously published criteria. In the absence of published criteria or a UKB-provided field for identifying those participants with a history of ADHD, ADHD cases were identified using ICD coding for ‘hyperkinetic disorder’ (ICD: F90). The ICD-10 uses the term ‘hyperkinetic disorders’ to describe early childhood-onset disorders characterised by impersistence with cognitive tasks and superfluous disorganised activity, rather than to describe ‘hyperkinetic movement disorders’ such as Huntington’s chorea. The codes for all phenotypes are detailed in the supplemental materials (Supplementary tables S6-S9).

#### UKB ‘controls’

Controls were samples that had neither epilepsy (Table 2) nor the psychiatric trait in question. We excluded from our controls those with HES codes or primary care codes suggestive of epilepsy or ASM use, those with mention of “seizures” or “convulsions” on their death certificate, those with self-reported ASM use, and those included in the UKB definition of epilepsy (data fields 131048 and 131049, Table 2).

**Table 2:** Identification of UK Biobank controls unlikely to have epilepsy.

| UK Biobank control cohort definition |  |
| --- | --- |
| In addition to not meeting our diagnostic definitions for epilepsy, psychosis, depression, anxiety, or ADHD cohorts, controls were selected from those UKB participants with primary care records available for interrogation and meeting none of the following criteria: |  |
| HES inpatient ICD-9 and ICD-10 codes | Codes indicating febrile or afebrile seizures or ASM (including acetazolamide, gabapentin, and pregabalin) |
| Primary care Read 2 and Read 3 diagnostic codes | Codes indicating febrile or afebrile seizures or ASM use (including acetazolamide, gabapentin, and pregabalin) |
| Self-reported medication history | ASM use (including acetazolamide, gabapentin, and pregabalin) |
| Death certification | “seizure” or “convulsi*” listed as primary or contributing cause of death |

#### Treatment response and epilepsy in UKB

To examine the relationship between PRS and the severity of epilepsy phenotype, we identified strata within our UKB epilepsy cohort with markers of resistant and responsive treatment outcomes.

‘Resistant’ cases had either inpatient hospital admissions for epilepsy or repeated electroencephalographic examination continuing well beyond initial presentation, neurosurgical intervention likely to have been for epilepsy, ASM use likely to indicate drug-resistance, or reference to epilepsy, status epilepticus or sudden unexpected death in epilepsy as the primary cause of death.

‘Responsive’ cases had markers suggestive of epilepsy in long-term remission or under sufficient control to not require admission to hospital, cessation of driving, further electroencephalographic investigation, or a new antiseizure medication. Given the last criterion, this group was restricted to those individuals with primary care data available for scrutiny of ASM prescriptions.

The data available in the UKB precluded the use of the formal International League Against Epilepsy definition of drug resistance^31^ because we could not be sure of the indication for each prescribed or self-reported ASM, and because we were unable to validate that the ASM was correctly selected for the individual’s epilepsy type or to exclude adverse effects as a reason for cessation of an ASM. Thus, we considered only ASM other than acetazolamide, gabapentin, and pregabalin, which are often prescribed for indications other than epilepsy. It is conceivable that in a small number of cases an efficacious ASM may have been stopped for pregnancy or family planning. However, we assumed that each ASM prescribed for 6 months was likely to have been well tolerated and its subsequent cessation is likely to indicate inefficacy rather than intolerance. Given these assumptions, we did not presume that commencement of a third ASM indicates drug-resistance. Instead, we conservatively stipulated that three ASM should be taken concurrently, a fourth ASM commenced, or hospital admission for epilepsy be required to indicate medication resistance. Similarly, indications for neurosurgery were unavailable so we identified individuals with recorded procedures consistent with surgical treatment of epilepsy.

#### Good and bad outcomes for epilepsy treatment from primary care data

People living with epilepsy in the UK should have an annual review in primary care of their seizure control, epilepsy treatment, and how epilepsy affects their daily functioning and quality of life NICE Clinical Knowledge Summary (https://cks.nice.org.uk/topics/epilepsy/management/routine-epilepsy-review/, accessed 2021-05-04). Consequently, primary care records include Read codes for summary outcomes which we evaluated for validation of our stratification filters. For comparison of strata by good outcomes we compared the frequency of annotations with any of Read 2 codes “21260” and “212J.” (epilepsy resolved) and “667C.” (epilepsy control good), and in Read 3, the additional codes “XaFjh” (epilepsy control good), and “XaJBZ” (epilepsy does not limit activities). For comparison by poor outcomes, we considered any of Read 2 codes “667D.” (epilepsy control poor), and “667K.” (epilepsy limits activities), and in Read 3 the additional codes “XaFji” (epilepsy control poor), “XaJBY” (epilepsy limits activities), and “XaJB4” (epilepsy restricts employment).

The EpiPGX consortium is a European project to investigate biomarkers of treatment response to anti-seizure medications. Drug resistant epilepsy in EpiPGX was defined as ≥4 seizures per year over the 12 months preceding the latest data entry, despite adequate trials of ≥2 tolerated and appropriately chosen ASMs, whether as monotherapies or in combination. Drug-responsive epilepsy was defined as freedom from seizures for ≥12 months up to the latest recorded visit^6^. Assignment of ‘responsive’ or ‘non-responsive’ in the EpiPGX cohort was based on evaluation of at least one epilepsy specialist at the recruiting site^32^.

### Genotype data quality control

Quality control on the genotype data was conducted using PLINK 1.9 ^33^. SNPs were removed if any of the following applied: genotype rate <0.98, minor allele frequency <0.01, or Hardy-Weinberg equilibrium deviation p<1 x 10^−6^. Samples were removed if they had genotyping call rate <0.98. To identify genetic outliers sample genotypes were thinned for linkage disequilibrium using PLINK’s “indep-pairwise” function (1000,100,0.1) and genetic principal components (PCs) were calculated using FlashPCAR^34^. The top two genetic PCs were plotted and assessed to ensure homogeneity across phenotype cohorts (Supplementary Figure S1).

### PRS analysis

The PRS analysis pipeline followed guidelines recently published guidelines, where appropriate ^35^. PRS for each trait of interest were calculated using PRSice 2.3.1^36^. The following GWAS statistics were used to calculate PRS for each of the relevant traits: epilepsy^17^, schizophrenia^37^, depression^38^, anxiety^39^, and ADHD^40^. As a negative control, PRS for rheumatoid arthritis^41^ were calculated and compared between epilepsy cases and controls (Supplementary figure S2). SNPs with p-values ≤0.5 from these GWAS were included in the PRS calculation. To verify p≤0.5 was an appropriate threshold for SNP inclusion in PRS modelling, we tested PRS models built from SNPs across a range of p-thresholds (Supplementary figure S3). Statistical analyses of the data were carried out in R v3.5^42^. PRS were normalised across all samples to mean 0 and standard deviation 1 and regressed onto phenotypes. The ggplot2 R package^43^ was used to graph the mean PRS and 95% confidence interval for all PRS.

The multcomp R package^44^ was used to estimate a multinomial linear regression model for all PRS calculated, with sex and the top eight genetic principal components included as covariates. As an additional control against population stratification affecting PRS results, we also included each sample’s North-South and East-West birth coordinates (as found in UKB phenotype fields f129 and f130, respectively, and normalised to mean 0 and SD 1) as covariates in all regression analysis (See Supplemental Data; Supplementary Tables S10 and S11).

We measured the prediction accuracy of PRS on a cohort of all UKB epilepsy samples (n=7 006). We conducted area under the receiver-operator curve analysis (AUC/ROC) with the pROC R package^45^, using PRS for each phenotype tested. Sex and the top eight PCs were included as covariates in the AUC/ROC analysis. To test the cumulative power of all PRS calculated in predicting epilepsy status, we created an additional model containing all PRS calculated combined, along with the top four PCs and sex as covariates (Supplementary Figure S4).

#### PRS in drug-resistant and responsive epilepsies

We examined the ability of psychiatric PRS to predict drug treatment of epilepsy using data from both the UKB and the EpiPGX Consortium. The phenotype criteria for ‘responsive’ epilepsy are provided above and in the supplements. PRS for each psychiatric trait of interest were calculated separately in EpiPGX and the UKB following the protocol described above. Binomial regression was used to compare PRS in resistant and responsive cases, with the top eight PCs and sex included as covariates. Each psychiatric PRS was incorporated into a fixed-effects meta-analysis model across the UKB and EpiPGX, using the ‘metafor’ R package ^46^

### Data availability

The data that support the findings of this study are available upon successful project application from the UK Biobank, and from the EpiPGX consortium.

## Results

### Cohort description

Of the 502 493 individuals from the UKB considered for this study, we identified 8 488 individuals with epilepsy. Following genotype quality control and screening for Caucasian ancestry, this figure fell to 409 634 individuals, which included 6 579 people with epilepsy. Control screening identified 286 502 samples with soft markers of epilepsy: these individuals were excluded from all analysis. Table 3 indicates the numbers of cases and controls included in each PRS analysis. Control numbers varied between analyses due to phenotype-specific control screening criteria.

**Table 3:** Cohort descriptions.

| Phenotype | Psych. phenotype only | Psych. phenotype AND epilepsy | Epilepsy only (no psych. phenotype) | Controls |
| --- | --- | --- | --- | --- |
| **Epilepsy** | - | - | 6 579 | 173 652 |
| **Psychosis** | 2 165 | 69 | 6 690 | 172 679 |
| **Depression** | 135 262 | 3 045 | 3 714 | 135 262 |
| **ADHD** | 82 | 3 | 6 757 | 179 617 |
| **Anxiety** | 16 990 | 507 | 6 252 | 166 153 |

**Table legend:** Indicated are the post-quality control numbers of samples for each phenotype tested. Control numbers vary due to the differing number of cases for each phenotype (i.e., controls in one set of analyses could be cases in another).

We defined two exclusive strata of epilepsy severity in the UKB: ‘Resistant’ and ‘Responsive’ (see Methods, Table 1). When compared to individuals in the responsive group, those in the resistant group were more likely to have poor outcomes codes and less likely to have good outcome codes (Table 4). Additionally, while there was no difference in age at onset, as might be expected from the inclusion criteria the Resistant group tended to have a greater duration of documented epilepsy, to report taking more ASM at enrolment, to have received more prescriptions for ASM, and to have received prescriptions for a greater number of distinct ASM (Supplementary Tables S1-5).

**Table 4:**
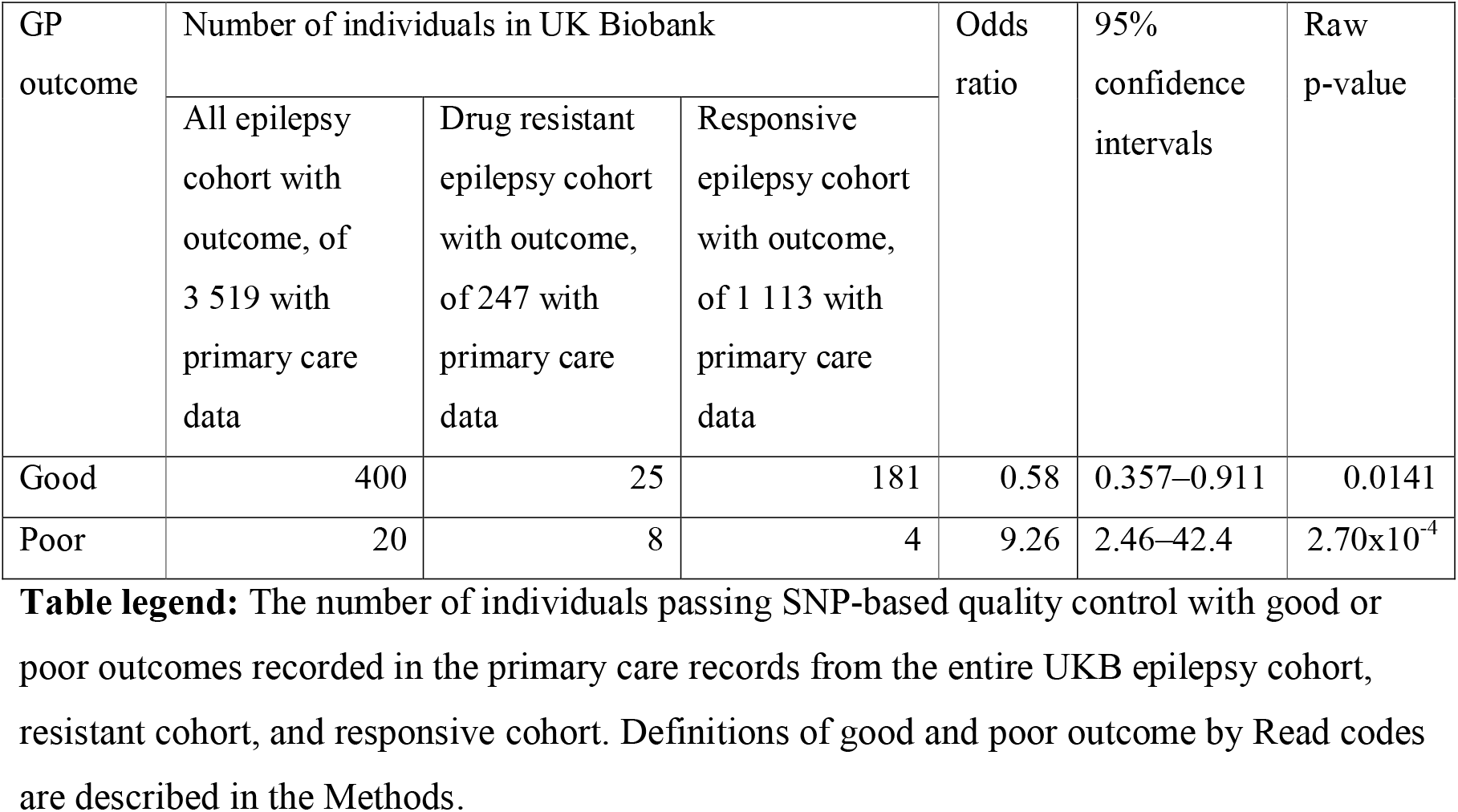
Comparison of outcome codes in primary care records by epilepsy severity strata.

### Epilepsy PRS as a predictor of epilepsy in UK Biobank

To test for enrichment of epilepsy PRS in epilepsy cases, we calculated and compared PRS using three separate epilepsy GWAS; ‘all epilepsy’, ‘focal epilepsy’ and ‘genetic generalised epilepsy’ (GGE) ^17^. We observed an elevation in epilepsy PRS in epilepsy cases relative to controls for all three PRS analyses (Figure 1). We found that ‘all epilepsy’ PRS explained the greatest phenotypic variance in case:control status (R^2^=0.23%), followed by GGE PRS (R^2^=0.13%) and focal PRS (R^2^=0.06%). Previously, PRS for GGE had been shown to differentiate GGE cases (defined by ICD coding) from controls in the UKB^20^. The results shown here, based on our reassessment of epilepsy phenotypes from a variety of UKB data sources, show significant enrichment of ‘all epilepsy’ and ‘focal epilepsy’ PRS in the UKB.

**Figure 1:**
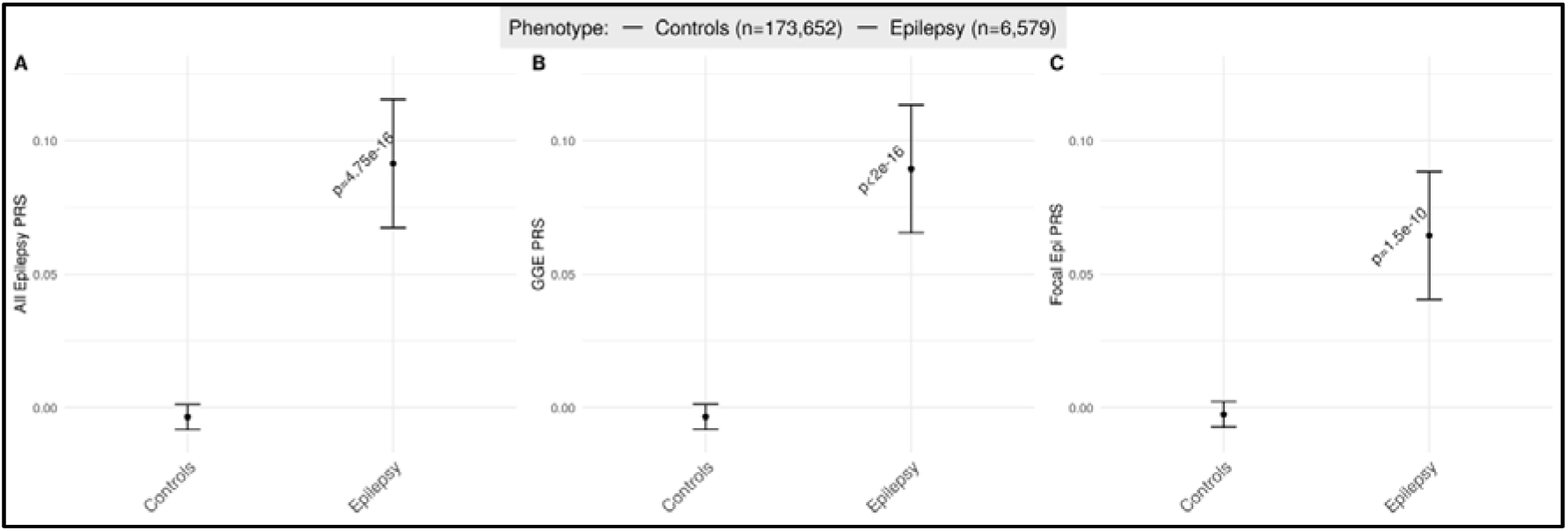
Epilepsy PRS is higher in UK Biobank epilepsy cases, relative to controls. PRS for A: All epilepsy, B: GGE, and C: Focal epilepsy. The y-axis shows the mean epilepsy PRS, error bars show 95% confidence intervals.

### Psychiatric PRS enrichment in epilepsy samples in the UK Biobank

#### Psychosis

To examine the role of schizophrenia PRS in epilepsy, we calculated and compared schizophrenia PRS across 1) people with epilepsy but no history of psychosis (n=6 690), 2) people with both epilepsy and psychosis (n=69), 3) people with psychosis but no history of epilepsy (n=2 165), and 4) population controls, who we suspect to have neither (n=172 679). We found that people with epilepsy and no history of psychosis had an elevated PRS for psychosis relative to the general population (p-corrected=0.014), and a lower PRS for psychosis than those with psychosis and no history of epilepsy (p-corrected<0.001) (Figure 2A). We confirmed that schizophrenia PRS is elevated in people with psychosis and no epilepsy, as previously described^30^. No significant results were found in any comparisons using the group with both psychosis and epilepsy (n=69).

**Figure 2:**
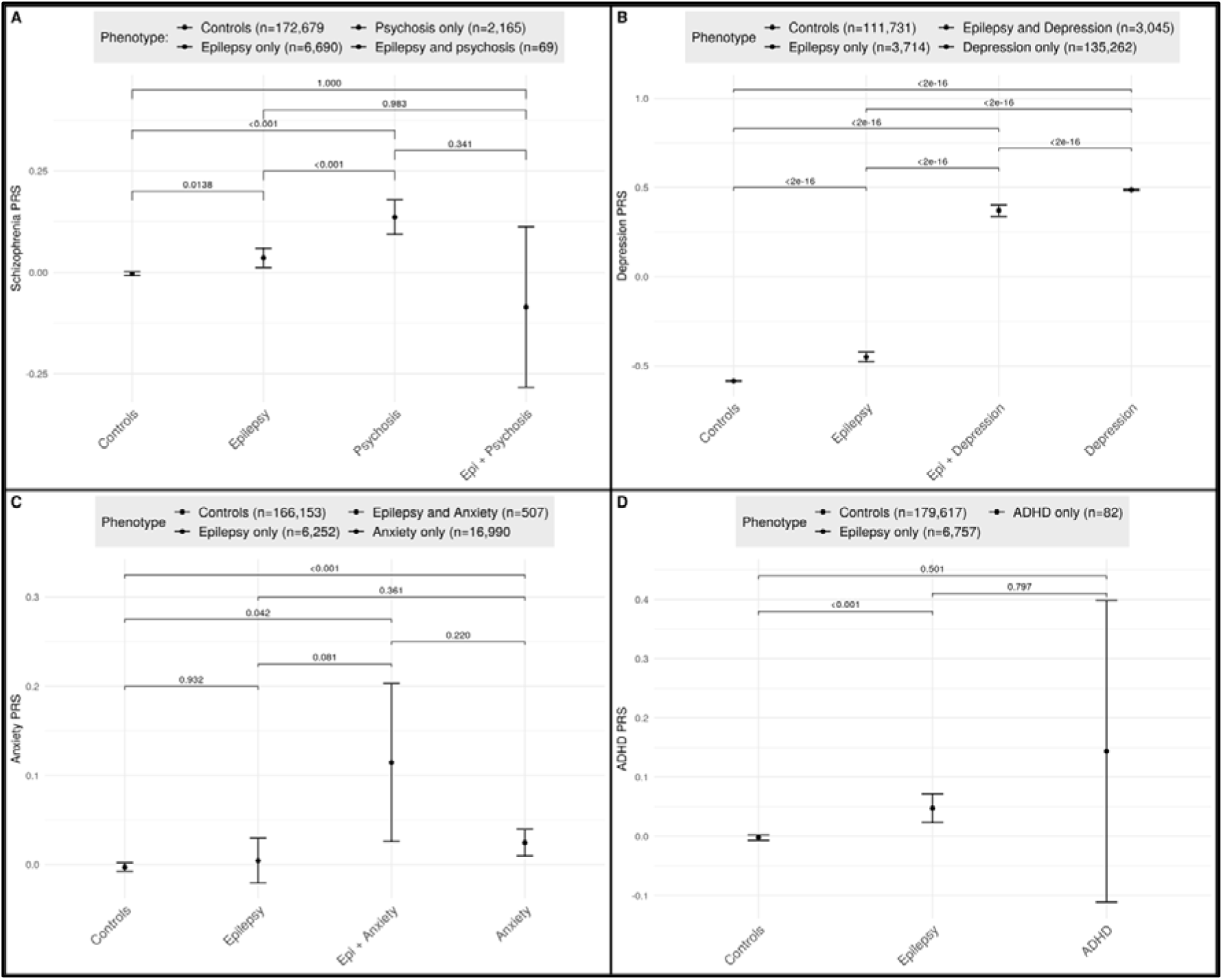
Psychiatric PRS analysis in UK Biobank samples with epilepsy and / or psychiatric disorders. PRS analyses are shown for A: Psychosis, B: Depression, C: Anxiety, D: ADHD. Phenotypes are displayed on the x-axes, normalised PRS on the y-axes. P-values displayed above brackets have been corrected for each pairwise comparison. Means and 95% CIs are displayed.

#### Depression

We calculated and compared depression PRS across 1) people with epilepsy but no history of depression (n=3 714), 2) people with both epilepsy and depression (n=3 045), 3) people with depression but no history of epilepsy (n=140 289), and 4) population controls, who we suspect to have neither (n=111 731). Of the groups examined, we observed the highest PRS for depression in those with depression only, followed by people with both epilepsy and depression (Figure 2B). Relative to population controls we observed an enrichment of depression PRS in people with epilepsy (p<2 x 10^−16^), people with epilepsy and depression (p<2 x 10^−16^), and people with depression but no epilepsy (p<2 x 10^−16^).

#### Anxiety

Anxiety PRS were compared across 1) people with epilepsy but no history of anxiety (n=6 252), 2) people with both epilepsy and anxiety (n=507), 3) people with anxiety but no history of epilepsy (n=16 990), and 4) population controls (n=166 153). We observed an enrichment of anxiety PRS in samples with anxiety and no epilepsy, relative to population controls (p-corrected<0.001), and samples with both epilepsy and anxiety (p-corrected=0.04), we found no enrichment for anxiety PRS in samples with epilepsy and no anxiety (Figure 2C).

#### ADHD

ADHD PRS were calculated and compared between 1) people with epilepsy but no history of ADHD (n=6 757), 2) people with ADHD but no history of epilepsy (n=69), and 3) population controls (n=179 617). As only 3 samples with both ADHD and epilepsy were found, they were excluded from this analysis. ADHD PRS was increased in samples with epilepsy relative to population controls (p-corrected=0.0001), but not in samples which met the phenotypic criteria for ADHD, which had wide confidence intervals (p=0.5) (Figure 2D).

#### AUC/ROC modelling

Incorporating any of the PRS tested into area under the receiver-operator characteristic curve analyses offered a modest increase in predictive ability over a null model (AUC range: 0.535 – 0.5483). Building a model comprised of all PRS tested, along with sex and the top 8 PCs provided a small, nonsignificant increase in AUC, indicating a likely correlation in the common genetic contributions to epilepsy status between the conditions tested (AUC: 0.5587) (Supplementary Figure S3).

### PRS differentiation across drug resistant and responsive cases of epilepsy

We were able to further phenotype the UKB epilepsy samples into 1 075 which were likely ‘responsive’ epilepsy, and 505 with likely ‘resistant’ epilepsy (see Table 1). We calculated and compared PRS for ‘all epilepsy’, ‘GGE’ and ‘focal epilepsy’ across these two treatment response categories (Figure 3). As three tests were conducted, the threshold for statistical significance was set at 0.0167. We found no significant difference of PRS for ‘all epilepsy’ (p=0.034), ‘focal epilepsy’ (p=0.0173) PRS, or GGE PRS (p=0.3372) between resistant and responsive cases. Relative to controls, ‘all epilepsy’ and ‘GGE’ PRS are higher in both responsive and resistant cases, while ‘focal epilepsy’ PRS is higher in resistant cases, but not responsive cases (Supplementary Table S12). We speculate that this is due to an enrichment of cases of focal epilepsy in our resistant cases relative to our responsive cases. However, the phenotype data available to us in the UKB were insufficient to explore this. As all samples from the EpiPGX dataset were included in the ILAE epilepsy GWAS^17^, and sample overlap is a known source of bias for PRS analysis^47^, we were unable to calculate epilepsy PRS in the EpiPGX samples.

**Figure 3:**
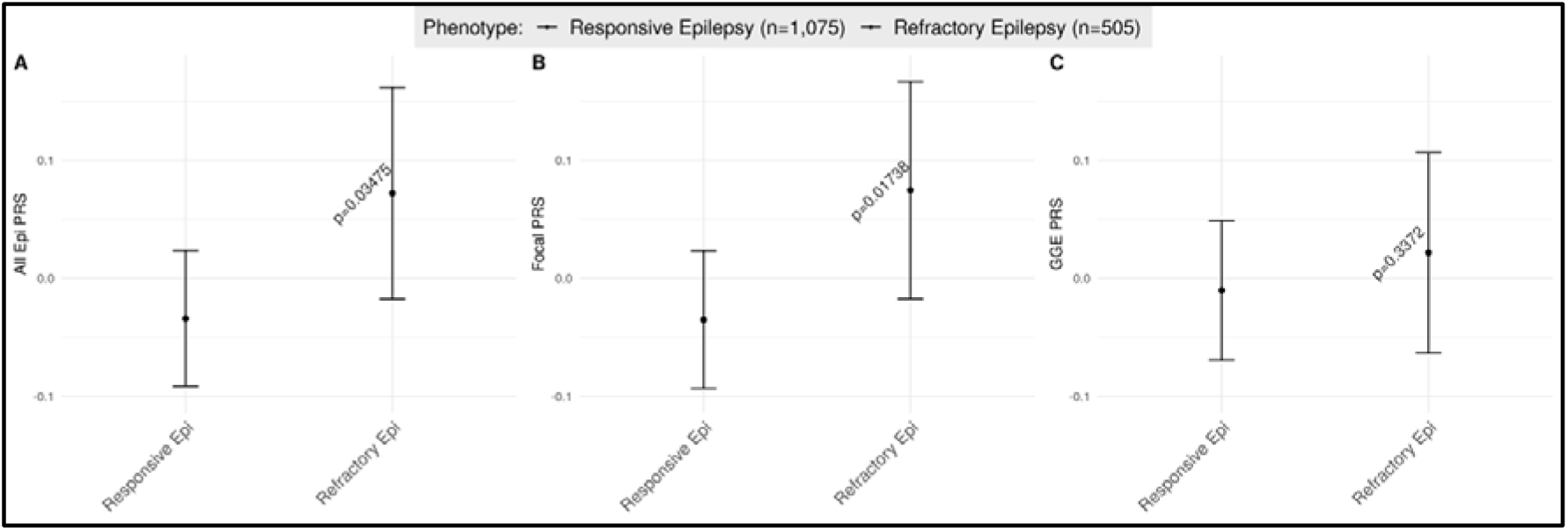
Epilepsy PRS comparison in UK Biobank responsive and resistant epilepsy cases. PRS for A: All epilepsy, B: Focal epilepsy, and C: GGEs. The y-axis shows the mean epilepsy PRS, error bars show 95% confidence intervals. P-values have been adjusted for covariates.

#### Psychiatric PRS in resistant and responsive epilepsy cases

Given the smaller numbers of samples of resistant and responsive samples available in the UKB, we opted to perform a fixed-effects meta-analysis analysis of psychiatric PRS in our UKB samples with samples obtained from the EpiPGX consortium. The EpiPGX epilepsy samples have been well-phenotyped by contributing clinical epileptologists according to treatment response allowing us to add data from 1 232 additional cases of responsive epilepsy and 2 217 cases of resistant epilepsy to these analyses.

We calculated PRS for each psychiatric trait of interest, used binomial regression to compare responsive and resistant epilepsies in EpiPGX and UKB separately and meta-analysed across the datasets. As 4 tests were carried out the threshold for significance was set at p<0.0125. We found higher PRS in resistant cases relative to controls for PRS developed for schizophrenia (p=0.0025) and depression (p=0.0008), but not ADHD (p=0.04) or anxiety (p=0.35) (Figure 4).

**Figure 4:**
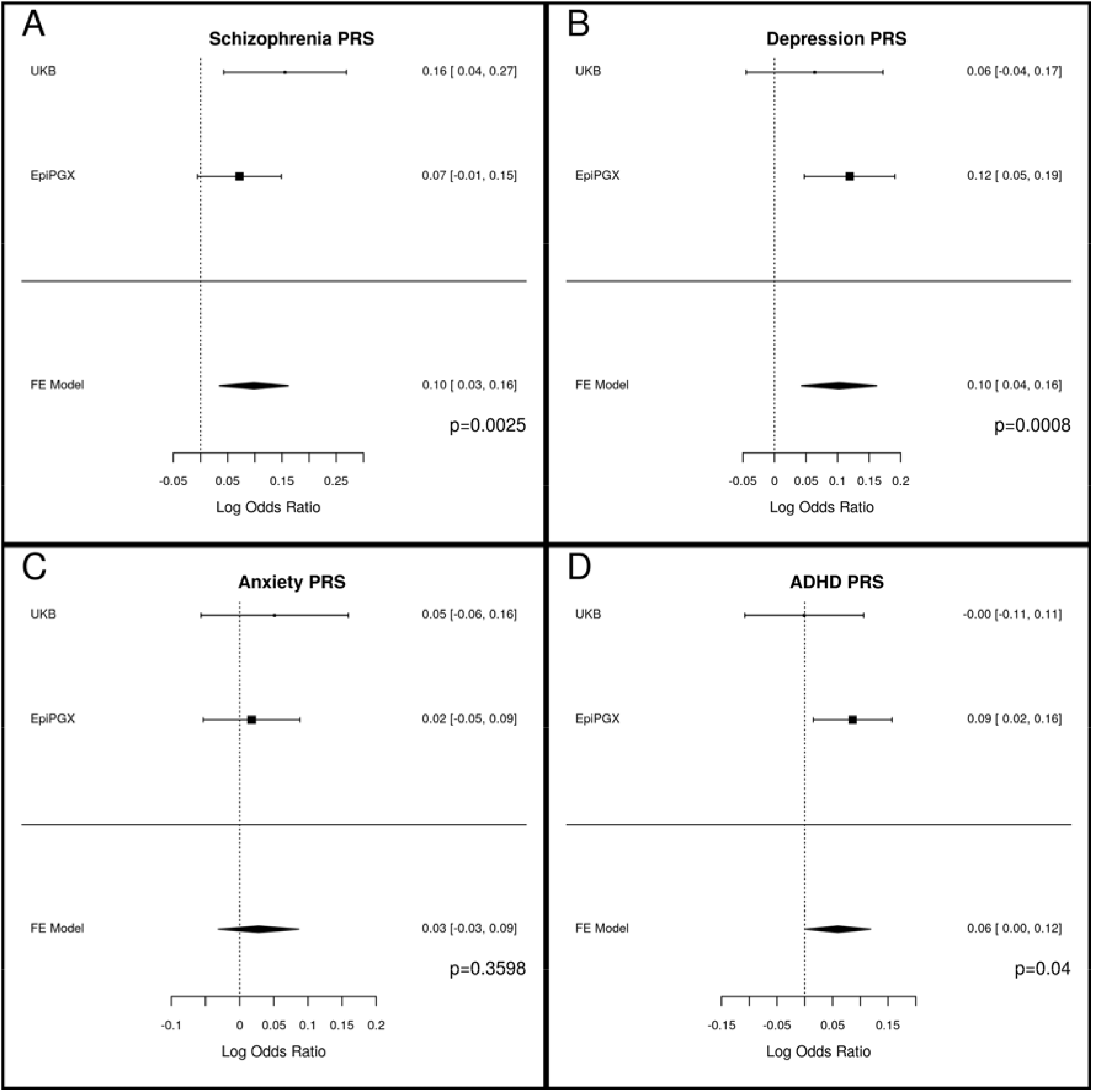
Fixed-effects meta-analysis of psychiatric PRS in cases of resistant and responsive epilepsy. PRS shown for A: psychosis, B: depression, C: anxiety, and D: ADHD. Odds ratios and standard errors for each PRS model in the UKB and EpiPGX datasets, and from a fixed-effects meta-analysis (FE model) of both datasets are displayed.

## Discussion

We aimed to test whether PRS for a variety of psychiatric conditions differed in people with common epilepsy relative to controls without epilepsy. We found that people with epilepsy have elevated PRS for depression, psychosis, and ADHD. We did not find an elevation for anxiety PRS in people with epilepsy. This presents the first evidence linking the common genetic basis of psychiatric conditions to a person’s risk of epilepsy and its responsiveness to treatment.

Previous studies looking for shared genetic links between epilepsy and common genetic comorbidities, using either LDSC^17^ or PRS^48^ did not identify any shared polygenic signals between epilepsy and its common psychiatric comorbidities^15^. In the study of Gui and colleagues from Hong Kong, the lack of genetic overlap could reflect limited study power for discovery (n=522 epilepsy cases, n=377 schizophrenia cases). Another potential explanation for these negative results is that PRS derived from GWAS of one population, may not be as predictive of a trait or condition in samples of a different genetic background^20, 49^. Gui and colleagues used PRS derived from mostly Caucasian samples and applied them to a cohort of mostly East Asian samples, which may reduce the predictive ability of the PRS. In contrast, others have shown that both PRS and LDSC were able to find genetic links shared among regularly co-presenting psychiatric phenotypes (Brainstorm Consortium 2018; Calafato *et al.* 2018). Our positive results here are likely to be at least partially attributable to i) the availability of recently published large GWAS for epilepsy and its various psychiatric comorbidities allowing for more robust PRS calculation, and ii) the relatively large number of people with epilepsy and controls, selected after detailed phenotypic exploration, available in the UKB allowing for even modest PRS signals to be detected.

Previous research^20^ has shown that epilepsy PRS is less suited to identifying cases of epilepsy from biobanks compared to cases from epilepsy-specific datasets comprised of samples from tertiary referral centres. This could potentially be due to the relatively mild epilepsy phenotypes of people in broader population databases compared to people with epilepsy attending tertiary referral centres, who may have a greater genetic burden for epilepsy. Alternatively, the threshold for inclusion in an epilepsy study is higher for those selected from tertiary centres by clinical academics than those included based on a self-declared or routine clinical diagnosis recorded in a biobank. Consequently, in a disorder with a significant rate of false positive diagnosis^51^, cohorts from tertiary centres are likely to be enriched for greater diagnostic certainty. Our findings demonstrate the value of detailed phenotypic data recently added to the UKB, allowing us to use the broader range of phenotypic sources to identify cases more critically than was previously possible for the study of Leu *et al.* (2019).

For depression we found that people with both epilepsy and depression had a lower depression PRS than people with depression and no epilepsy. From these results we hypothesise that epilepsy is acting as an additional ‘environmental’ burden for depression, meaning that the presence of epilepsy and its consequences is sufficient to precipitate depression in people who are at lower genetic risk. Clinical observations would corroborate the idea that epilepsy can act as an environmental burden for depression. Certain anticonvulsants, including levetiracetam and topiramate, are known to cause or exacerbate mood disorders^52^, and epilepsy patients who undergo successful epilepsy surgery show long-term mood improvements^53^. We also speculate that a similar pattern is true for the other conditions, however due to smaller numbers of samples in these cohorts, and potentially as their PRS are derived from less robust GWAS, we were unable to detect such a pattern with any degree of confidence.

In this study we had insufficient cases to test the role of ADHD PRS in a similar manner to which we tested the other psychiatric traits, given that we only identified 82 cases of ADHD (via the proxy phenotype coding for ‘hyperkinetic disorder’), and 3 cases with both ADHD and epilepsy. Larger cohort sizes, and cohorts in younger populations in which ADHD is more likely to have been diagnosed^54^, would provide more clarity to the aetiological links between epilepsy and ADHD. We also had smaller numbers of individuals with psychosis than would be expected based on population estimates, whereas our sample numbers for depression and anxiety were closer to expectations^9^, highlighting the variability of disease phenotype availability and potential recruitment bases of the UKB, which is known not to be a cohort representative of the general UK population.

A potential confounder to our results is that cases included in the various psychiatric GWAS used to calculate PRS were not screened for epilepsy, and as such the GWAS may contain cryptic associations with epilepsy. However, controls from the psychiatric GWAS were also not screened for epilepsy status, which would somewhat lessen the potential enrichment of genetic signals associated with epilepsy. We note that our phenotyping of epilepsy cases was unable to distinguish confidently between cases of focal and generalised epilepsy or particular syndromes. Future work could aim to delineate genetic burden for psychiatric illness based on epilepsy type.

Incorporating PRS into predictive models using AUC/ROC analysis showed modest predictive ability. Previous work comparing PRS analysis from population datasets to disease-specific studies found a stronger PRS signal in the disease-specific studies ^55^. This presents a significant challenge to the development of PRS as a tool to identify disease cases from the general population. More research in this area is needed, and the development of such methods was beyond the scope of this study.

This research also found for the first time higher PRS burden for psychosis and depression in drug resistant relative to responsive epilepsies. These increased PRS correlate with higher rates of psychiatric disorders in resistant epilepsy relative to responsive epilepsy^11^. The increased PRS observed in the resistant cohorts could indicate that common genetic variants that drive psychiatric illness also drive difficult-to-treat seizures, or that common genetics variants which are associated with psychiatric illness can drive pharmacoresistance in individuals with epilepsy.

Environmental factors and the consequences of chronic seizures on quality of life may also be contributing to the increased incidence of psychiatric illness in drug resistant epilepsy. Other neuro-biological factors may further drive the incidence of psychiatric illness in resistant epilepsies. For example, the prevalence of both drug resistant seizures and psychiatric illness is high in people with temporal lobe epilepsies^56^, however genetic factors may confound this.

These results show that common genetic signals associated with a variety of psychiatric conditions are also enriched in people with epilepsy. This has implications for understanding of genetic aetiology of epilepsy, and its relevant psychiatric comorbidities. The results shown here could be potentially useful for prognostic stratification of epilepsy, based on multiple PRS, and identification of those with epilepsy who are predisposed to psychiatric comorbidities. This work motivates an examination of the interplay between PRS and rare variants which are known to contribute to both seizure disorders and psychiatric disorders.

## Data Availability

All data produced are available online through an application to UK Biobank (https://www.ukbiobank.ac.uk/)

## Acknowledgements

This project has been conducted using the UK Biobank resource under project ID 35124. This research was funded in part, by the Wellcome Trust [203914/Z/16/Z] and by Science Foundation Ireland (SFI) under Grant Number 16/RC/3948 and co-funded under the European Regional Development Fund and by FutureNeuro industry partners. For the purpose of open access, the author has applied a CC BY public copyright licence to any Author Accepted Manuscript version arising from this submission. We thank the participants of the UK Biobank and the people with epilepsy who contributed their data to this study.

## Supplementary material

Supplementary material is available online

## References

1. Ngugi AK, Bottomley C, Kleinschmidt I, Sander JW, Newton CR. Estimation of the burden of active and life-time epilepsy: a meta-analytic approach. Epilepsia. 2010 May;51(5):883–90.

2. Beghi E, Giussani G, Nichols E, Abd-Allah F, Abdela J, Abdelalim A, et al. Global, regional, and national burden of epilepsy, 1990&#x2013;2016: a systematic analysis for the Global Burden of Disease Study 2016. Lancet Neurol [Internet]. 2019 Apr 1;18(4):357–75. Available from: https://doi.org/10.1016/S1474-4422(18)30454-X

3. Myers KA, Johnstone DL, Dyment DA. Epilepsy genetics: Current knowledge, applications, and future directions. Clin Genet. 2019 Jan;95(1):95–111.

4. Balestrini S, Arzimanoglou A, Blümcke I, Scheffer IE, Wiebe S, Zelano J, et al. The aetiologies of epilepsy. Epileptic Disord. 2021 Feb;23(1):1–16.

5. Goldenberg MM. Overview of drugs used for epilepsy and seizures: etiology, diagnosis, and treatment. P T. 2010 Jul;35(7):392–415.

6. Chen Z, Brodie MJ, Liew D, Kwan P. Treatment Outcomes in Patients With Newly Diagnosed Epilepsy Treated With Established and New Antiepileptic Drugs: A 30-Year Longitudinal Cohort Study. JAMA Neurol. 2018 Mar;75(3):279–86.

7. Tellez-Zenteno JF, Patten SB, Jette N, Williams J, Wiebe S. Psychiatric comorbidity in epilepsy: a population-based analysis. Epilepsia. 2007 Dec;48(12):2336–44.

8. Kanner AM, Ribot R, Mazarati A. Bidirectional relations among common psychiatric and neurologic comorbidities and epilepsy: Do they have an impact on the course of the seizure disorder? Epilepsia open. 2018 Dec;3(Suppl Suppl 2):210–9.

9. Campbell C, Cavalleri GL, Delanty N. Exploring the genetic overlap between psychiatric illness and epilepsy: A review. Epilepsy Behav. 2020 Jan;102:106669.

10. Clarke MC, Tanskanen A, Huttunen MO, Clancy M, Cotter DR, Cannon M. Evidence for Shared Susceptibility to Epilepsy and Psychosis: A Population-Based Family Study. Biol Psychiatry [Internet]. 2017 Aug 25;71(9):836–9. Available from: http://dx.doi.org/10.1016/j.biopsych.2012.01.011

11. Hellwig S, Mamalis P, Feige B, Schulze-Bonhage A, van Elst LT. Psychiatric comorbidity in patients with pharmacoresistant focal epilepsy and psychiatric outcome after epilepsy surgery. Epilepsy Behav. 2012 Mar;23(3):272–9.

12. Eaton CB, Thomas RH, Hamandi K, Payne GC, Kerr MP, Linden DEJ, et al. Epilepsy and seizures in young people with 22q11.2 deletion syndrome: Prevalence and links with other neurodevelopmental disorders. Epilepsia [Internet]. 2019/04/11. 2019 May;60(5):818–29. Available from: https://www.ncbi.nlm.nih.gov/pubmed/30977115

13. Kolc KL, Sadleir LG, Scheffer IE, Ivancevic A, Roberts R, Gecz J. A systematic review and meta-analysis of 271 PCDH19-variant individuals identi fi es psychiatric comorbidities, and association of seizure onset and disease severity. Mol Psychiatry [Internet]. 2019;241–51. Available from: http://dx.doi.org/10.1038/s41380-018-0066-9

14. Knott S, Forty L, Craddock N, Thomas RH. Epilepsy and bipolar disorder. Epilepsy Behav. 2015 Nov;52(Pt A):267–74.

15. Brainstorm Consortium, Anttila V, Bulik-Sullivan B, Finucane HK, Walters RK, Bras J, et al. Analysis of shared heritability in common disorders of the brain. Science [Internet]. 2018 Jun 22;360(6395):eaap8757. Available from: https://www.ncbi.nlm.nih.gov/pubmed/29930110

16. Bulik-Sullivan BK, Loh P-R, Finucane HK, Ripke S, Yang J, Consortium SWG of the PG, et al. LD Score regression distinguishes confounding from polygenicity in genome-wide association studies. Nat Genet. 2015/02/02. 2015 Mar;47(3):291–5.

17. ILAE. Genome-wide mega-analysis identifies 16 loci and highlights diverse biological mechanisms in the common epilepsies. Nat Commun. 2018 Dec;9(1):5269.

18. van Rheenen W, Peyrot WJ, Schork AJ, Lee SH, Wray NR. Genetic correlations of polygenic disease traits: from theory to practice. Nat Rev Genet. 2019 Oct;20(10):567– 81.

19. Ripke S, Dushlaine CO, Chambert K, Moran JL, Anna K, Akterin S, et al. Genome-wide association analysis identifies 13 new risk loci for schizophrenia. Nat Genet [Internet]. 2013;45(10):1–26. Available from: http://www.nature.com/ng/journal/v45/n10/abs/ng.2742.html

20. Leu C, Stevelink R, Smith AW, Goleva SB, Kanai M, Ferguson L, et al. Polygenic burden in focal and generalized epilepsies. Brain [Internet]. 2019 Oct 14;142(11):3473–81. Available from: https://doi.org/10.1093/brain/awz292

21. Bulik-Sullivan B, Finucane HK, Anttila V, Gusev A, Day FR, Loh P-R, et al. An atlas of genetic correlations across human diseases and traits. Nat Genet. 2015 Nov;47(11):1236–41.

22. Pouget JG, Han B, Wu Y, Mignot E, Ollila HM, Barker J, et al. Cross-disorder analysis of schizophrenia and 19 immune-mediated diseases identifies shared genetic risk. Hum Mol Genet. 2019 Oct;28(20):3498–513.

23. Markota M, Coombes BJ, Larrabee BR, McElroy SL, Bond DJ, Veldic M, et al. Association of schizophrenia polygenic risk score with manic and depressive psychosis in bipolar disorder. Transl Psychiatry [Internet]. 2018;8(1):188. Available from: https://doi.org/10.1038/s41398-018-0242-3

24. Leu C, Richardson TG, Kaufmann T, van der Meer D, Andreassen OA, Westlye LT, et al. Pleiotropy of polygenic factors associated with focal and generalized epilepsy in the general population. PLoS One. 2020;15(4):e0232292.

25. Bycroft C, Freeman C, Petkova D, Band G, Elliott LT, Sharp K, et al. The UK Biobank resource with deep phenotyping and genomic data. Nature [Internet]. 2018;562(7726):203–9. Available from: https://doi.org/10.1038/s41586-018-0579-z

26. Fonferko-Shadrach B, Lacey AS, White CP, Powell HWR, Sawhney IMS, Lyons RA, et al. Validating epilepsy diagnoses in routinely collected data. Seizure. 2017 Nov;52:195–8.

27. Mbizvo GK, Bennett KH, Schnier C, Simpson CR, Duncan SE, Chin RFM. The accuracy of using administrative healthcare data to identify epilepsy cases: A systematic review of validation studies. Epilepsia. 2020 Jul;61(7):1319–35.

28. Purves KL, Coleman JRI, Meier SM, Rayner C, Katrina AS, Cheesman R, et al. A Major Role for Common Genetic Variation in Anxiety Disorders. 2019;

29. Howard DM, Adams MJ, Clarke T-K, Hafferty JD, Gibson J, Shirali M, et al. Genome-wide meta-analysis of depression identifies 102 independent variants and highlights the importance of the prefrontal brain regions. Nat Neurosci. 2019 Mar;22(3):343–52.

30. Legge SE, Jones HJ, Kendall KM, Pardinas AF, Menzies G, Bracher-Smith M, et al. Association of Genetic Liability to Psychotic Experiences With Neuropsychotic Disorders and Traits. JAMA psychiatry. 2019 Sep;

31. Kwan P, Arzimanoglou A, Berg AT, Brodie MJ, Allen Hauser W, Mathern G, et al. Definition of drug resistant epilepsy: consensus proposal by the ad hoc Task Force of the ILAE Commission on Therapeutic Strategies. Epilepsia. 2010 Jun;51(6):1069–77.

32. Wolking S, Schulz H, Nies AT, McCormack M, Schaeffeler E, Auce P, et al. Pharmacoresponse in genetic generalized epilepsy: a genome-wide association study. Pharmacogenomics. 2020 Apr;21(5):325–35.

33. Chang CC, Chow CC, Tellier LC, Vattikuti S, Purcell SM, Lee JJ. Second-generation PLINK: rising to the challenge of larger and richer datasets. Gigascience. 2015;4:7.

34. Abraham G, Qiu Y, Inouye M. FlashPCA2: principal component analysis of Biobank-scale genotype datasets. Bioinformatics. 2017 Sep;33(17):2776–8.

35. Wand H, Lambert SA, Tamburro C, Iacocca MA, O’Sullivan JW, Sillari C, et al. Improving reporting standards for polygenic scores in risk prediction studies. Nature [Internet]. 2021;591(7849):211–9. Available from: https://doi.org/10.1038/s41586-021-03243-6

36. Choi SW, O’Reilly PF. PRSice-2: Polygenic Risk Score software for biobank-scale data. Gigascience [Internet]. 2019 Jul 15;8(7). Available from: https://doi.org/10.1093/gigascience/giz082

37. Ripke S, Neale BM, Corvin A, Walters JTR, Farh K-H, Holmans PA, et al. Biological Insights From 108 Schizophrenia-Associated Genetic Loci. Nature [Internet]. 2014 Jul 24;511(7510):421–7. Available from: http://www.ncbi.nlm.nih.gov/pmc/articles/PMC4112379/

38. Wray NR, Eriksson N, Escott-Price V, Farhadi F, Kiadeh H, Finucane HK. Genome-wide association analyses identify 44 risk variants and refine the genetic architecture of major depressive disorder. 2017; Available from: https://www.biorxiv.org/content/biorxiv/early/2017/07/24/167577.full.pdf

39. Otowa T, Hek K, Lee M, Byrne EM, Mirza SS, Nivard MG, et al. Meta-analysis of genome-wide association studies of anxiety disorders. Mol Psychiatry [Internet]. 2016 Jan 12;21:1391. Available from: http://dx.doi.org/10.1038/mp.2015.197

40. Savage JE, Jansen PR, Stringer S, Watanabe K, Bryois J, de Leeuw CA, et al. Genome-wide association meta-analysis in 269,867 individuals identifies new genetic and functional links to intelligence. Nat Genet. 2018 Jul;50(7):912–9.

41. Okada Y, Wu D, Trynka G, Raj T, Terao C, Ikari K, et al. Genetics of rheumatoid arthritis contributes to biology and drug discovery. Nature [Internet]. 2013/12/25. 2014 Feb 20;506(7488):376–81. Available from: https://pubmed.ncbi.nlm.nih.gov/24390342

42. R Core Team. R: A Language and Environment for Statistical Computing [Internet]. Vienna, Austria; 2019. Available from: https://www.r-project.org/

43. Wickham H. ggplot2: Elegant Graphics for Data Analysis [Internet]. Springer-Verlag New York; 2016. Available from: https://ggplot2.tidyverse.org

44. Hothorn T, Bretz F, Westfall P. Simultaneous Inference in General Parametric Models. Biometrical J. 2008;50(3):346–63.

45. Robin X, Turck N, Hainard A, Tiberti N, Lisacek F, Sanchez J-C, et al. pROC: an open-source package for R and S+ to analyze and compare ROC curves. BMC Bioinformatics. 2011;12:77.

46. Viechtbauer W. Conducting Meta-Analyses in R with the metafor Package. J Stat Software; Vol 1, Issue 3 [Internet]. 2010; Available from: https://www.jstatsoft.org/v036/i03

47. Wray NR, Yang J, Hayes BJ, Price AL, Goddard ME, Visscher PM. Pitfalls of predicting complex traits from SNPs. Vol. 14, Nature reviews. Genetics. 2013. p. 507–15.

48. Gui H, Li M, Sham PC, Baum L, Kwan P, Cherny SS. Genetic overlap between epilepsy and schizophrenia: Evidence from cross phenotype analysis in Hong Kong Chinese population. Am J Med Genet B Neuropsychiatr Genet. 2018 Jan;177(1):86– 92.

49. Martin AR, Gignoux CR, Walters RK, Wojcik GL, Neale BM, Gravel S, et al. Human Demographic History Impacts Genetic Risk Prediction across Diverse Populations. Am J Hum Genet [Internet]. 2017;100(4):635–49. Available from: http://dx.doi.org/10.1016/j.ajhg.2017.03.004

50. Calafato MS, Thygesen JH, Ranlund S, Zartaloudi E, Cahn W, Crespo-Facorro B, et al. Use of schizophrenia and bipolar disorder polygenic risk scores to identify psychotic disorders. Br J Psychiatry [Internet]. 2018 Sep;213(3):535–41. Available from: https://pubmed.ncbi.nlm.nih.gov/30113282

51. Xu Y, Nguyen D, Mohamed A, Carcel C, Li Q, Kutlubaev MA, et al. Frequency of a false positive diagnosis of epilepsy: A systematic review of observational studies. Seizure [Internet]. 2016;41:167–74. Available from: https://www.sciencedirect.com/science/article/pii/S1059131116301339

52. Elger CE, Johnston SA, Hoppe C. Diagnosing and treating depression in epilepsy. Seizure. 2017 Jan;44:184–93.

53. Hamid H, Liu H, Cong X, Devinsky O, Berg AT, Vickrey BG, et al. Long-term association between seizure outcome and depression after resective epilepsy surgery. Neurology [Internet]. 2011 Nov 29;77(22):1972 LP – 1976. Available from: http://n.neurology.org/content/77/22/1972.abstract

54. Chung W, Jiang S-F, Paksarian D, Nikolaidis A, Castellanos FX, Merikangas KR, et al. Trends in the Prevalence and Incidence of Attention-Deficit/Hyperactivity Disorder Among Adults and Children of Different Racial and Ethnic Groups. JAMA Netw open. 2019 Nov;2(11):e1914344.

55. Oetjens MT, Kelly MA, Sturm AC, Martin CL, Ledbetter DH. Quantifying the polygenic contribution to variable expressivity in eleven rare genetic disorders. Nat Commun [Internet]. 2019;10(1):4897. Available from: https://doi.org/10.1038/s41467-019-12869-0

56. de Oliveira GNM, Kummer A, Salgado JV, Portela EJ, Sousa-Pereira SR, David AS, et al. Psychiatric disorders in temporal lobe epilepsy: an overview from a tertiary service in Brazil. Seizure. 2010 Oct;19(8):479–84.

