## Supplemental data for "Polygenic risk score analysis reveals shared genetic burden between epilepsy and psychiatric comorbidities"

The below tables show the phenotypic criteria used to define UKB cohorts beyond epilepsy (see Table 1 in the main manuscript) for this study**:**

| Psychosis | | |
|---|---|---|
| **UK Biobank field** | **Field name** | **Code** |
| 20471 | Ever seen an un-real vision | Yes |
| 20463 | Ever heard an un-real voice | Yes |
| 20474 | Ever believed in un-real voices or signs | Yes |
| 20468 | Ever believed in an un-real conspiracy against self | Yes |
| 20462 | Distress caused by unusual or psychotic experiences | A bit distressing / Quite distressing / Very distressing (4,5,6) |

**Table S6:** Psychosis: phenotype criteria for 'distressing psychotic experiences' were taken from(Legge *et al.* 2019).

| Depression | | |
|---|---|---|
| **UK Biobank Field** | **Field name** | **Code** |
| 2090 | Ever seen a GP for nerves, anxiety, tension, or depression? | Yes |
| 2010 | Ever seen a psychiatrist for anxiety, tension, nerves, or depression | Yes |
| 41202 | ICD 10 (main) | F32 / F33 / F34 / F38 / F39 (Depressive episode / Recurrent Depressive disorder / Persistent mood disorders / Other mood disorders / Unspecified mood disorders) |
| 41203 | ICD 10 (secondary) | F32 / F33 / F34 / F38 / F39 |

**Table S7:** Depression: Phenotype for 'Broad Depression' taken from (Howard *et al.* 2019).

| ADHD | | |
|---|---|---|
| **UK Biobank Field** | **Field name** | **Code** |
| 41202 | ICD 10 (main) | F90 (Hyperkinetic disorder) |
| 41203 | ICD 10 (secondary) | F90 (Hyperkinetic disorder) |

**Table S8:** ADHD, using Hyperkinetic disorder as a surrogate.

| Anxiety | | |
|---|---|---|
| **UK Biobank field** | **Field name** | **Code** |
| 20544 | Mental health problems ever diagnosed by a professional | Social anxiety / phobia / panic attacks / anxiety / agoraphobia (1,3,6,15,17) |
| 20421 | Ever felt worried, tense, or anxious for a month or longer | Yes |
| 20420 | Longest time spent anxious | >1 month |
| 20538 | Worried most days during period of worst anxiety | Yes |
| 20540 | Multiple worries during worst period of anxiety | Yes |
| 20543 | Number of things worried about during worst period of anxiety | >1 |
| 20541 | Difficulty stopping worrying during worst period of anxiety | Yes |
| 20539 | Frequency of inability to stop worrying during worst period of anxiety | Sometimes/often |
| 20537 | Frequency of difficulty controlling worry during worst period of anxiety | Sometimes/often |
| 20429 | Easily tired during worst period of anxiety | Yes |
| 20419 | Difficulty concentrating during worst period of anxiety | Yes |
| 20422 | More irritable than usual during worst period of anxiety | Yes |
| 20417 | Tense, sore, or aching muscles during worst period of anxiety | Yes |
| 20427 | Frequent trouble falling or staying asleep during worst period of anxiety | Yes |
| 20418 | Impact on normal roles during worst period of anxiety | Somewhat / a lot |

**Table S9:** Anxiety: Phenotype definitions taken from (Purves *et al.* 2019) for "probable lifetime generalised anxiety disorder".

**Principal component analyses (PCA):**

2D PCA was used to ensure that no genetic outliers were included in any of our analyses.

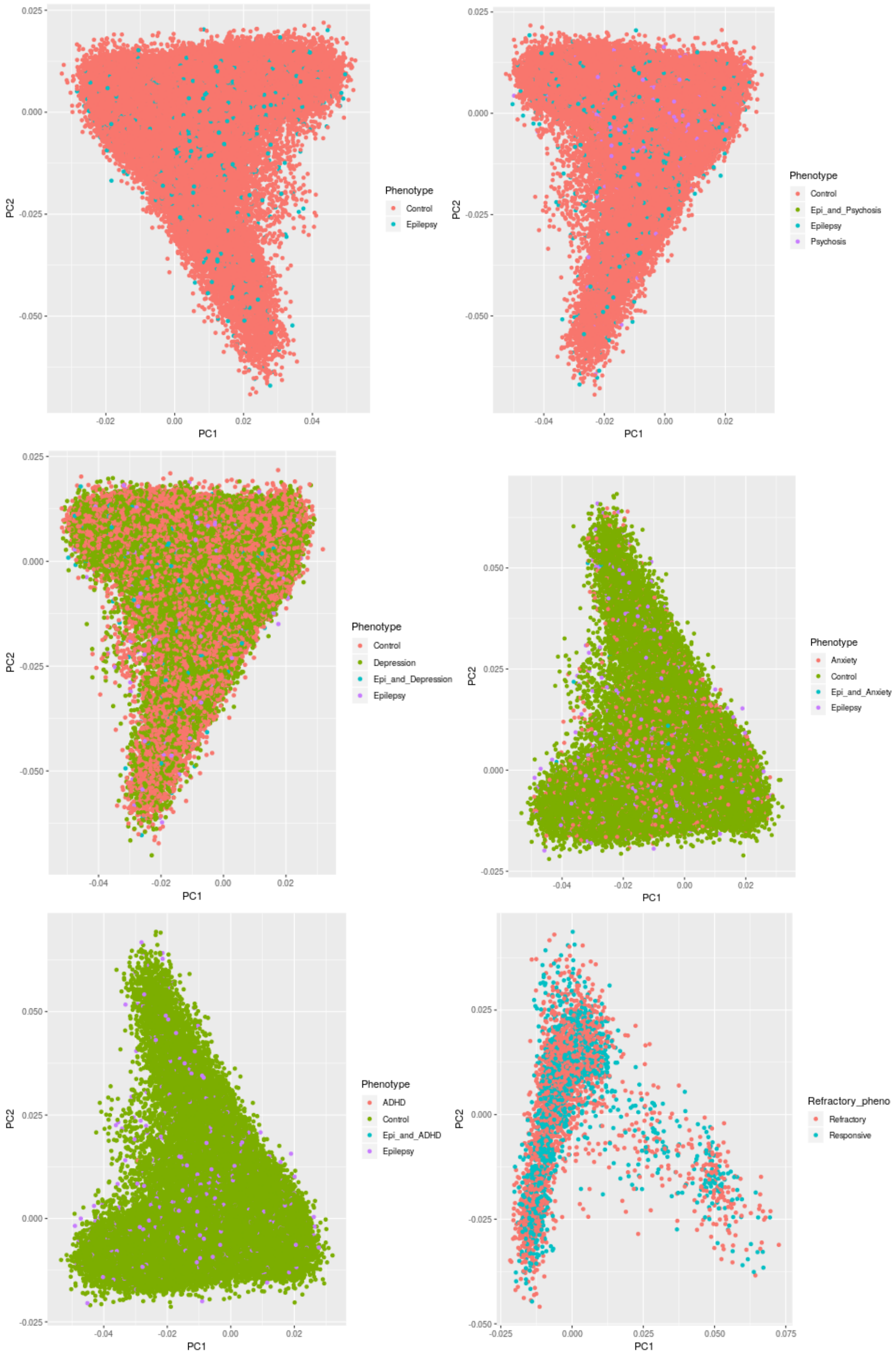

**Supplementary Figure S1:** Showing PCA for each cohort analysed. Phenotypes denoted by colour.

**Control PRS:**

To verify that the differences seen in any of the above PRS were not being driven by any confounding factors, such as population stratification, we calculated PRS for rheumatoid arthritis (RA; Okada et al., 2014) and compared between epilepsy cases (n=7 006) and controls (n=179 736). The PRS was calculated by taking SNPs from a variety of p-thresholds in the RA GWAS. No significant differences were detected at any p-threshold tested.

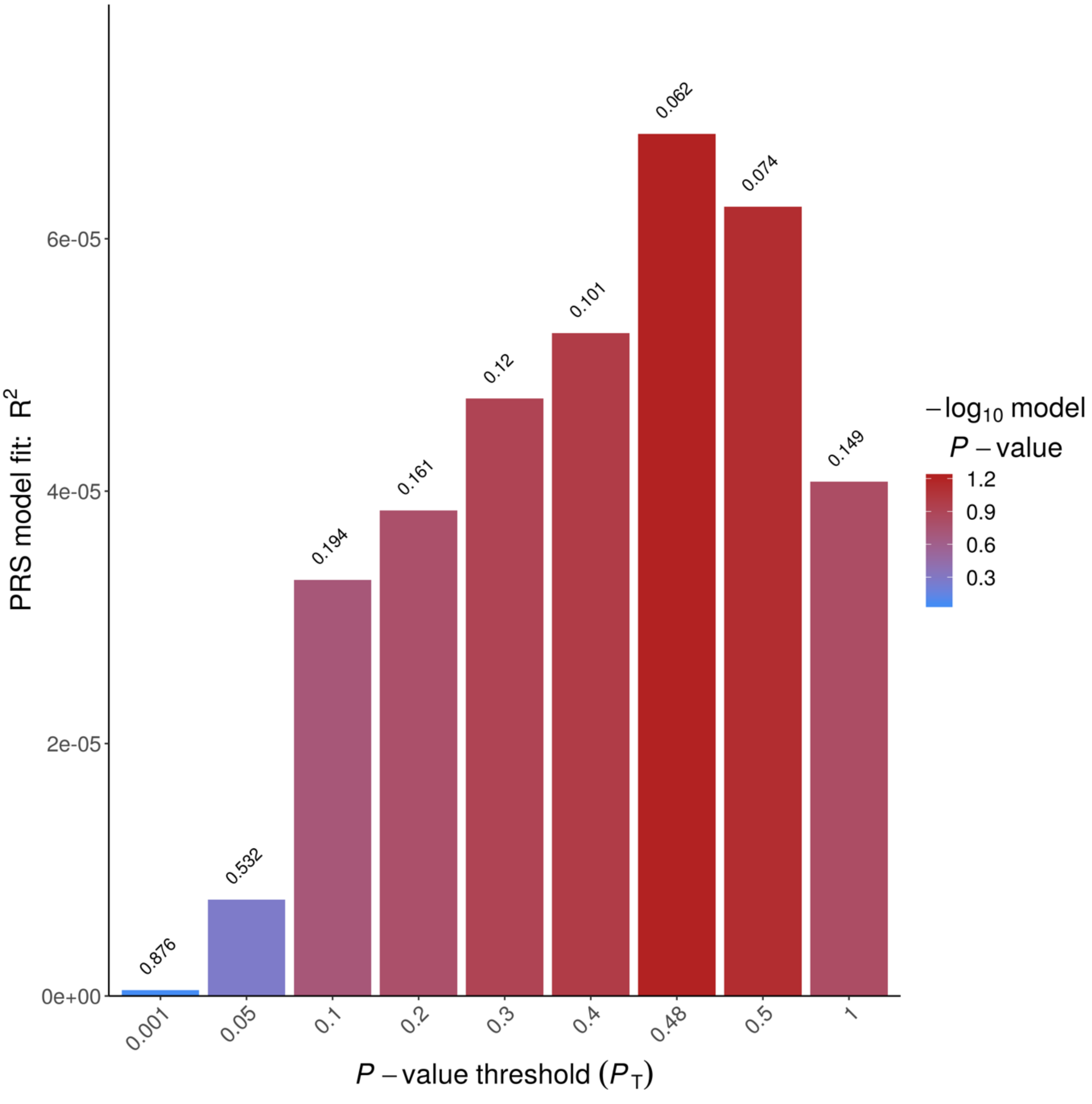

**Supplementary Figure S2:** Showing the significance levels of arthritis PRS models comparing epilepsy cases and controls. X-axis denotes the p-threshold used to calculate PRS, y-axis shows $R^2$ of PRS model at each threshold. Numbers above bars and bar colours show p-values. The p-threshold which was closest to significance was $P_T$=0.48 (p=0.062).

**Thresholding:**

In order to verify that $P_T$=0.5 was an appropriate p-threshold to use for PRS modelling, we calculated PRS models comparing cases and controls for each of the traits we studied across 10 values of $P_T$ using PRSICE2, including the top 4 PCs and sex as covariates. For each PRS model, $P_T$=0.5 was either the most significant $P_{T,}$ or explained a near-identical proportion of the phenotypic variance as the most significantly associated $P_T$.

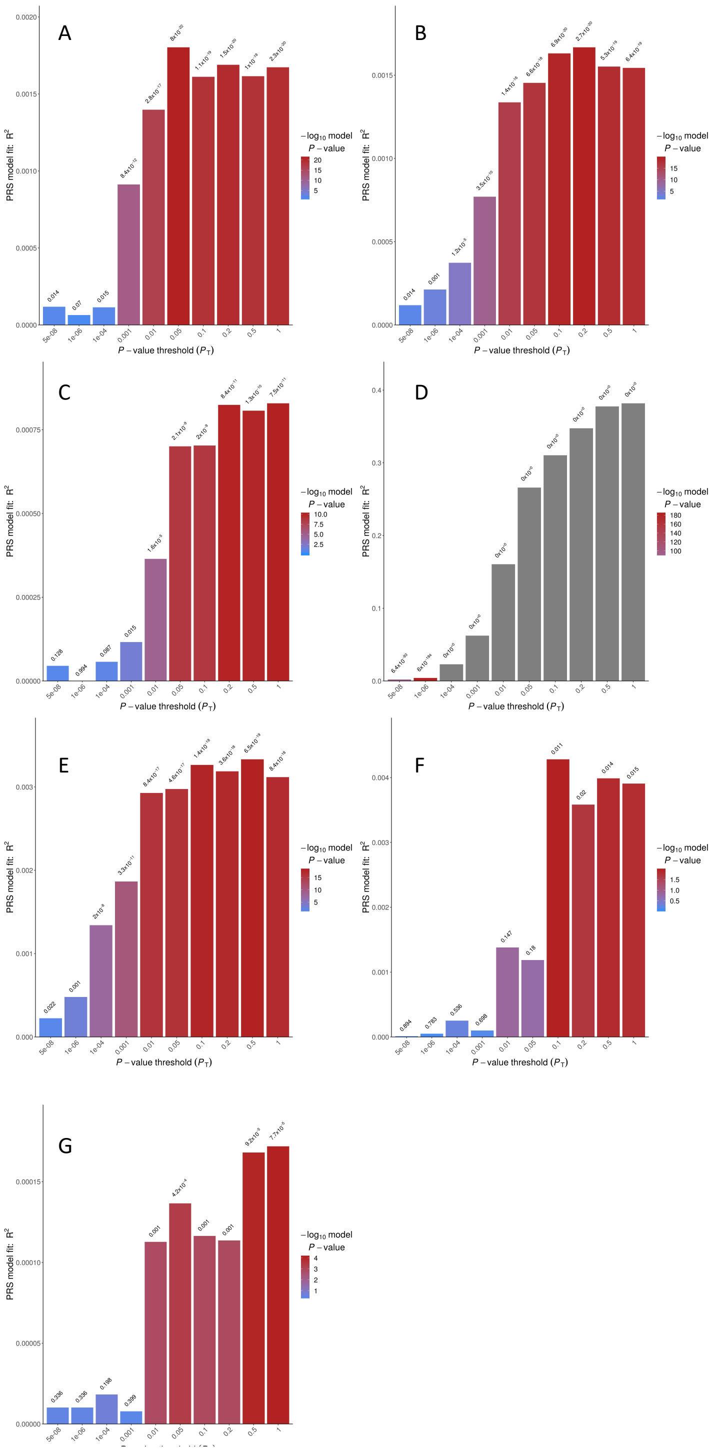

**Supplementary Figure S3:** Showing PRS models for A) all epilepsy, B) GGE, C) Focal epilepsy, D) Depression, E) Schizophrenia, F) ADHD, and G) Anxiety across a range of $P_T$s. The x-axis displays each $P_T$s , the y-axis shows the variance explained by each PRS model. Bar colours and numbers above each bar show significance.

**Controlling for Population stratification:**

To further verify that our PRS models are not solely driven by ancestral differences between samples, we compared the birth location of samples in our cohorts to PRS. We extracted North-South (NS) and East-West (EW) birth coordinates for all available samples in our cohorts using UKB phenotype fields f129 and f130, respectively. These coordinates were then normalised to mean 0 and SD1 using the scale() function in R. We ran linear models comparing each PRS to birth coordinates. We then ran logistic modelling regressing phenotype onto PRS, NS/EW coordinates, PCs 1-4, and sex.

For most PRS we observed either the north-south or east-west coordinate was significantly associated with PRS, but incorporating birth location coordinates into ANOVA modelling comparing PRS to phenotype and covariates did not change which PRS were significantly associating with phenotype.

| **All Epilepsy** | | | |
|---|---|---|---|
| | Estimate | Std.Error | P |
| **(Intercept)** | -3.181256 | 0.017769 | $<2^{-16}$ |
| **Allepi_PRS** | 0.106221 | 0.013117 | 5.59E-16 |
| **NS** | -0.261085 | 0.01645 | $<2^{-16}$ |
| **EW** | -0.034163 | 0.014545 | 0.0188 |
| **PC1** | 0.144538 | 0.01461 | $<2^{-16}$ |
| **PC2** | 0.094857 | 0.014916 | $2.03^{-10}$ |
| **PC3** | -0.009989 | 0.012457 | 0.4226 |
| **PC4** | -0.015076 | 0.012704 | 0.2354 |
| **as.factor(Sex)2** | -0.171755 | 0.024829 | 4.60E-12 |

**Table S10:** Showing results of binomial regression model comparing all epilepsy PRS and covariates between samples with epilepsy and controls.

| Depression PRS | | | | |
|---|---|---|---|---|
| | SumSq | MeanSq | Fvalue | P |
| **Phenotype** | 71540 | 23847 | 34258 | $<2^{-16}$ |
| **PC1** | 100 | 100 | 143.33 | $<2^{-16}$ |
| **PC2** | 39 | 39 | 56.05 | $7.09^{-14}$ |
| **PC3** | 483 | 483 | 693.22 | $<2^{-16}$ |
| **PC4** | 3247 | 3247 | 4665.06 | $<2^{-16}$ |
| **NS** | 91 | 91 | 130.25 | $<2^{-16}$ |
| **EW** | 18 | 18 | 25.58 | $4.25^{-07}$ |
| **as.factor(Sex)** | 1606 | 1606 | 2307.61 | $<2^{-16}$ |
| **Anxiety PRS** | | | | |
| | SumSq | MeanSq | Fvalue | P |
| **Phenotype** | 19 | 6.2 | 6.201 | 0.00033 |
| **PC1** | 598 | 598.4 | 601.473 | $<2^{-16}$ |
| **PC2** | 11 | 10.6 | 10.615 | 0.001122 |
| **PC3** | 324 | 324.3 | 325.962 | $<2^{-16}$ |
| **PC4** | 13 | 12.9 | 13.013 | 0.000309 |
| **NS** | 9 | 9.5 | 9.5 | 0.002056 |
| **EW** | 1 | 0.8 | 0.827 | 0.363135 |
| **as.factor(Sex)** | 1 | 1.5 | 1.484 | 0.223228 |
| **Schizophrenia PRS** | | | | |
| | SumSq | MeanSq | Fvalue | P |
| **Phenotype** | 51 | 17 | 18.447 | $5.83^{-12}$ |
| **PC1** | 7945 | 7945 | 8661.012 | $<2^{-16}$ |
| **PC2** | 3114 | 3114 | 3394.439 | $<2^{-16}$ |
| **PC3** | 1430 | 1430 | 1559.192 | $<2^{-16}$ |
| **PC4** | 2428 | 2428 | 2646.358 | $<2^{-16}$ |
| **NS** | 45 | 45 | 49.063 | $2.49^{-12}$ |
| **EW** | 0 | 0 | 0.478 | 0.489 |
| **as.factor(Sex)** | 2 | 2 | 2.208 | 0.137 |
| **ADHD PRS** | | | | |
| | SumSq | MeanSq | Fvalue | P |
| **Phenotype** | 19 | 6.2 | 6.252 | 0.000307 |
| **PC1** | 10 | 10.1 | 10.091 | 0.00149 |
| **PC2** | 8 | 7.7 | 7.683 | 0.005573 |
| **PC3** | 409 | 409.2 | 410.203 | $<2^{-16}$ |
| **PC4** | 0 | 0 | 0 | 0.997171 |
| **NS** | 0 | 0 | 0 | 0.987447 |
| **EW** | 7 | 6.5 | 6.562 | 0.010421 |

| | | | | |
|---|---|---|---|---|
| **as.factor(Sex)** | 2 | 2.5 | 2.485 | 0.114933 |

**Table S11**: Showing the full results of ANOVA testing comparing PRS to phenotype (control, epilepsy, epilepsy + psych issue), psych issue only, birth coordinates, and covariates. In all instances, PRS remains significantly associated with phenotype, despite also correlating with covariates and birth location. NS = North-South birth coordinate, EW = East-West birth coordinate, SumSq = sum of squares, MeanSq = Mean square (variance).

**AUC/ROC modelling:**

In order to determine the relative ability of each PRS to identify samples with epilepsy in the UKB, we calculated the area under the receiver-operator characteristic curve for each phenotype examined (see methods). We found that any PRS model tested had a higher AUC than the base model of sex + PCs only (Figure 3). The depression PRS was model was found to have a higher prediction accuracy than epilepsy PRS. This is likely potentially because 45% of epilepsy samples also have depression, and that the depression GWAS from which the PRS were calculated is currently more robust than that of epilepsy. Building a model comprised of all PRS tested, along with sex and the top 8 PCs was the most predictive model generated. However, the difference is marginal, and far from additive, which indicates that there is likely an overlap in the common genetic contributions to epilepsy status between the conditions tested.

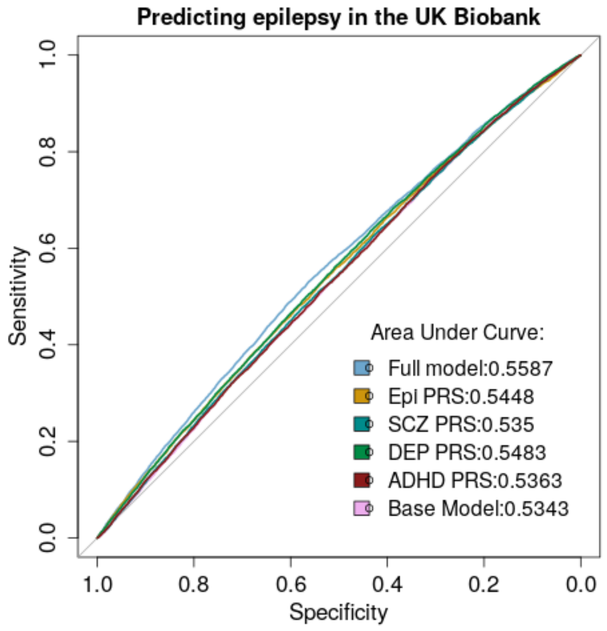

**Supplementary Figure 4:** AUC/ROC analysis on epilepsy samples in the UKB. The phenotypes being tested are shown in the bottom right corner. The 'full model' model is built from a regression of all PRS calculated, along with sex and the top 8 principal components. Epi = epilepsy, SCZ = schizophrenia, DEP = depression.

**Epilepsy PRS in drug resistant and responsive cases of epilepsy, relative to controls:**

We calculated and compared PRS for ‘all epilepsy’, ‘focal epilepsy’, and ‘GGE’ in the UKB, and compared both our resistant and responsive cases to controls. Shown below in table S12 are the uncorrected p-values of responsive and resistant epilepsy cases, relative to controls. As 6 tests were performed the threshold of significance is 0.0088.

| | **Responsive** | **Resistant** |
|---|---|---|
| **All epilepsy PRS** | $2.98^{-04}$ | $9.23^{-09}$ |
| **GGE PRS** | $7.40^{-05}$ | $1.09^{-05}$ |
| **Focal PRS** | $4.16^{-02}$ | $2.51^{-06}$ |

**Table S12:** Showing the p-values, relative to controls, of ‘all epilepsy’, ‘GGE’, and ‘focal epilepsy’ PRS in the UKB responsive and resistant cohorts. The p-values shown are uncorrected.
